# Cost-effectiveness analysis of the geko™ device (an NMES technology) in managing venous leg ulcers in UK healthcare setting

**DOI:** 10.1101/2024.06.13.24308720

**Authors:** Richard Tuson, Andrew Metry, Keith Harding

**Author notes:** **Corresponding author:** Richard Tuson, Health Analytical Solutions Ltd, Macclesfield, Cheshire, England –.

## Abstract

**Objective:** This study evaluates the cost-effectiveness of the geko device a neuromuscular electro-stimulator (NMES) technology with standard of care versus standard of care alone for venous leg ulcers treatment, from the UK National Health Service perspective over 12 months.

**Setting:** Research was conducted across NHS UK facilities, primarily within community services and outpatient leg ulcer clinics, encompassing a total of 51 patients.

**Method:** A partitioned survival model, based on a two-arm randomised controlled trial, assessed wound healing rates using Kaplan–Meier curves and parametric extrapolations over a 12-month time horizon. Costs were derived from UK reference costs the British National Formulary, and the Personal Social Services Research Unit (2021/22). The primary outcome measured was the incremental cost per quality-adjusted life-year gained. The geko device provides additional benefits by stimulating the lateral popliteal nerve, augmenting venous, arterial and microvascular flow.

**Results:** The addition of the geko device to standard of care significantly enhanced outcomes, increasing healing probability by 68% compared to standard of care. This integration would result in a cost saving of £774.14 per patient when compared to the standard of care alone across the NHS. Economic analyses indicate that integrating the geko device into standard of care protocols would reduce the overall NHS expenditure on venous leg ulcer wound management by as much as 15%. The approach also positively impacted health-related quality of life.

**Conclusion:** The geko™ device when used adjunctively with standard of care would be a cost-effective method for managing chronic venous leg ulcers within the NHS, improving healing rates and offering economic benefits.

**Key messages:** *Enhanced Healing Rates:* The addition of the geko™ device (an NMES technology) to standard of care significantly increases the probability of healing in patients with chronic venous leg ulcers.

*Economic Advantage:* Incorporating the geko device into standard of care would lead to a reduction in the overall costs of wound management in the NHS, potentially up to 15%.

*Cost-Effectiveness:* The combination of the geko device and standard of care demonstrates cost-effectiveness, offering a favourable balance between costs and quality-adjusted life years gained.

*Improved Quality of Life:* The use of the geko device in addition to standard of care impacts positively the health-related quality of life for patients.

*Consistent Outcomes in Sensitivity Analysis:* Sensitivity analyses support the cost dominance of the combined treatment approach, indicating robustness in various scenarios.

*Policy Implications:* These findings suggest that the integration of the geko device technology would be a viable policy option for enhancing venous leg ulcer treatment within the NHS.

## 1. Introduction

Venous leg ulcers (VLUs) represent a substantial public health challenge in the UK. They are estimated to affect approximately 1 in 500 individuals, a rate which increases to one in 50 for those over the age of 80 ^**(1)**^. These ulcers, primarily caused by inadequate venous return and often exacerbated by factors such as age, obesity, family history, and deep vein thrombosis, are a leading cause of morbidity and significantly impact health-related quality of life (HRQoL) (2,3). The National Wound Care Strategy (NWCS), initiated by NHS England in conjunction with the Academic Health Sciences Network, underscores the growing burden and economic impact of lower limb wound care, including VLUs. In 2019, the strategy identified around 739,000 cases of leg ulcers in England, with the associated healthcare costs amounting to an estimated £3.1 billion annually ^**(4)**^. This considerable expenditure and the report’s emphasis on the lack of effective, evidence-based care for many patients highlight the urgent need for improved treatment strategies. The strategy further suggests that without decisive action, the prevalence of such wounds could increase by up to 4% per year, highlighting the critical need for efficient and effective management approaches in the face of this escalating healthcare challenge.

As of February 2024, the NHS confronts a challenging scenario with an elective waiting list surpassing 7.5 million, bed occupancy rates exceeding 90% for elective and acute, and over 20,000 elective operations cancelled in the third quarter of 2023—averaging nearly 261 cancellations daily ^**(5)**^. A contributor to this strain is the admission of patients with worsening venous leg ulcers (VLUs).

It is reported in the UK that the burden cost of VLUs could be around the £5,488 per patients ^**(6)**^ and in comparison in other countries Germany reports a similar cost of €6,905 ^**(7)**^.

The healing process of VLUs varies greatly, with some ulcers healing within weeks, while others may take several months or even become chronic ^**(2)**^.

The two most important variables affecting the rate of healing of VLUs are known to be size of ulcer and duration of ulceration. Despite this most patients are treated only with compression systems ^**(8)**^

The geko™ device, a product of Firstkind Ltd, UK, is a neuromuscular electro-stimulator (NMES) technology designed for non-invasive use. It is applied to the skin’s surface on the lateral aspect of the leg below the knee and over the head of the fibula. The device delivers a balanced electrical pulse to the common peroneal nerve, inducing dorsiflexion of the foot, which activates the venous calf and foot muscle pumps, thereby enhancing venous, arterial and microvascular blood flow. In this paper, the term ‘the active arm’ specifically refers to the geko device used for 12 hours a day as an adjunct to standard of care (SoC). Also, ‘the control arm’ refers to standard of care which has been applied under the NHS facilities current VLU treatment protocol. This geko device has been classified as a class IIa medical device and has received a CE and UKCA mark.

In a previously reported study ^**(2)**^., a randomised controlled trial (RCT) examining the efficacy of the NMES technology was conducted with patients suffering from chronic VLUs in a UK setting. As depicted in the flow diagram (Figure 1) from the previous mentioned study ^**(2)**^, the cost-effectiveness comparison was conducted post-randomisation, separating the two arms. Data from this analysis, detailed in ^**(2)**^, guided the emphasis on this subgroup within the health economics evaluation. In our study, a total of 51 patients that were allocated post randomisation and compliant with the protocol of ^**(2)**^, was 29 receiving the active arm and 22 receiving the control arm alone. The demographic and wound characteristics were comparable across both groups at baseline. The VLUs were classified as chronic in a multidisciplinary clinic setting or by healthcare professionals if they remained unhealed for 4–6 weeks. At the 16-week mark, the previously report RCT study ^**(2)**^ reported healing rates of 42% for the active arm and 27% for the control arm. Using Kaplan–Meier analysis, these rates were projected to 12-month healing rates of 0.84 for the active arm and 0.50 for the control arm alone, equating to mean healing times of 25.3 weeks and 37.6 weeks, respectively. The mean wound bed area was 9.961 cm^2^ for the active arm and 10.390 cm^2^ for the control arm. Additionally, an improvement in pain scores from baseline to the end of treatment was observed, being 30.1% for the active arm and 21.1% for the control arm.

**Figure 1.**
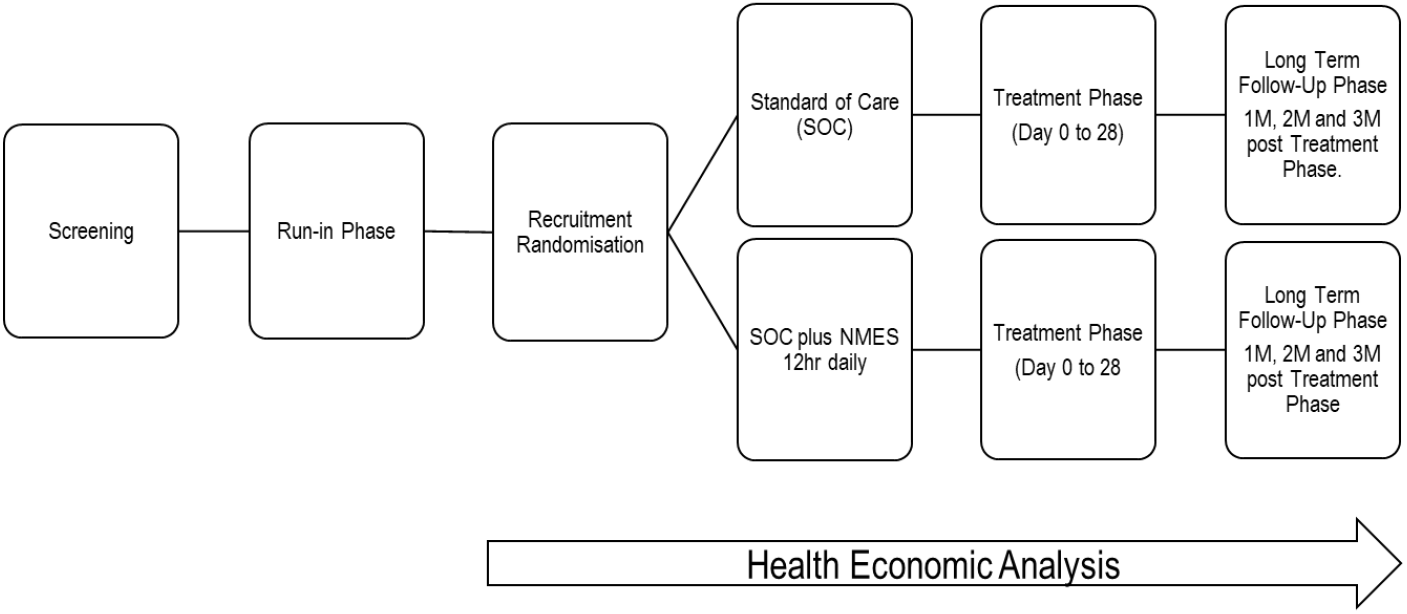
Trial schematic detailing health economic analysis period. The healing trajectory of VLUs is highly variable; some ulcers may resolve within weeks, while others may take several months or years to heal ^**(9)**^. Chronic VLUs are characterised by prolonged inflammation and often lead to recurring infections, requiring extensive clinical management and representing a significant burden on healthcare resources ^**(10)**^.

## Method

### 2.1 Design of the Cost-Effectiveness Analysis

In this economic evaluation, we conducted a modelling study based on a two-arm, RCT ^**(2)**^. Our approach involved cohort analysis, utilising anonymised patient records of individuals diagnosed with VLUs within the UK healthcare system. A key focus of this study is to equip clinicians with a nuanced understanding of the economic aspects of VLU treatments, particularly in relation to the NMES technology, which we evaluate as a potential cost-saving alternative to the current SoC in the NHS. The primary objective is to assess the cost-effectiveness and potential economic benefits of various treatment strategies and interventions for VLUs. This research not only examines the clinical efficacy of the active arm but also highlights its economic impact, especially on healthcare budgets. By integrating current cost data and examining outcomes in real-world settings, our analysis provides crucial insights into the financial dimensions of managing chronic VLUs in a dynamically evolving healthcare environment, facilitating decisions that balance clinical excellence with fiscal responsibility.

### 2.2 Economic Modelling

We developed a partitioned survival model to depict the management of chronic VLUs using Microsoft Excel (Figure 2.). This model considered both clinical costs and outcomes in relation to the choice of treating a VLU with the active arm or the control arm. The modelling spanned a 12-month time horizon. Initially, chronic VLUs were introduced into the model in an open state, with two treatment pathways: the active arm or the control arm. Progression within the model involved transitioning to one of three health states: open (ulcer remaining untreated), infected (ulcer clinically diagnosed with an infection), or closed (ulcer completely healed) ^**(11)**^. The ulcers could either persist in their current state or transition weekly to a different state over the course of the year. It is important to note that a VLU could occupy only one health state at any given time within the model’s timeframe, as the health states were selected due to them being mutually exclusive ^**(12)**^.

**Figure 2.**
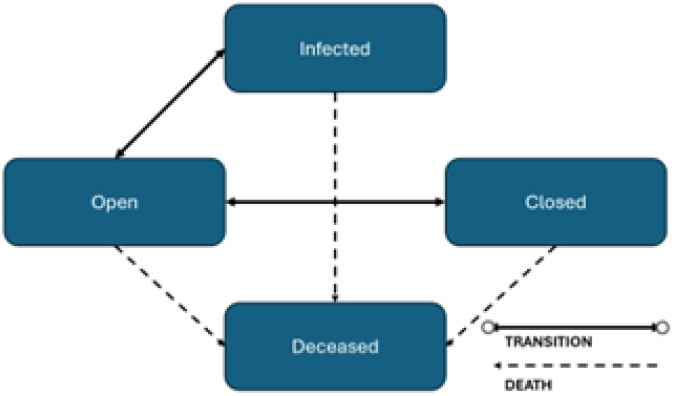
Partitioned survival model.

Data populating this model included transition probabilities, clinical outcomes, resource usage, and utility estimates. Given the original study protocol’s duration of 16 weeks, it was necessary to extend these findings to a 12-month period. This extension was accomplished using Kaplan– Meier survival curve data (Figures 3,4), which provided the basis for the healing trajectory (^**13**^). The extrapolation employed six parametric models: Weibull, exponential, Gompertz, log-logistic, log-normal, and generalised gamma distributions. Following this extrapolation, clinicians conducted a visual review to determine the most accurate fit, drawing on their experience with both the active arm and the control arm. Professor Keith Harding, an experienced figure in VLU research in the UK, verified the appropriateness of the chosen parametric extrapolations, endorsing the Weibull model for the active arm and the log-logistic model for the control arm.

**Figure 3.**
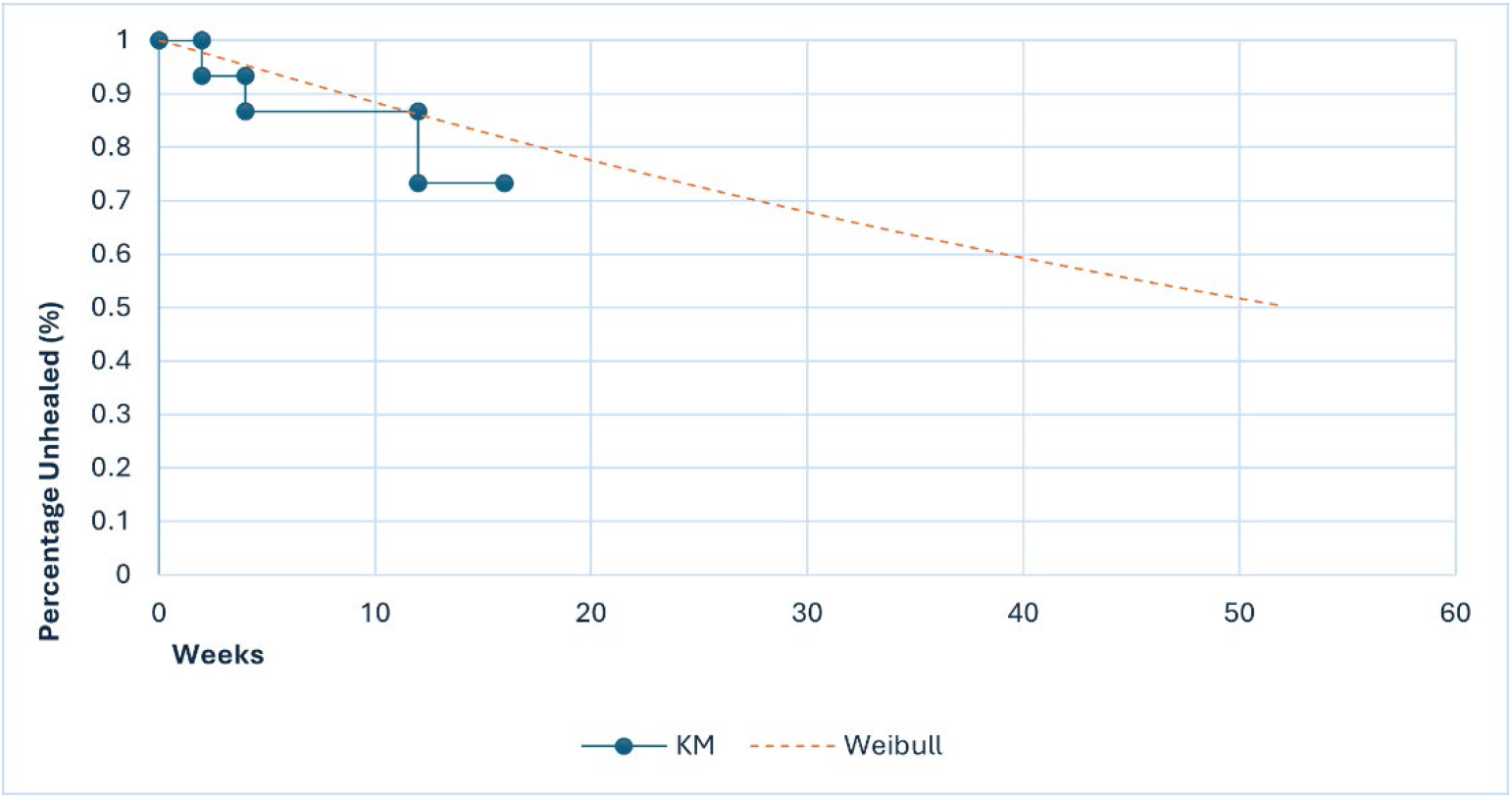
Kaplan–Meier (KM) time-to-healing analysis with parametric extrapolations for the control arm showing Weibull distribution. Control Arm, NHS SoC only.

**Figure 4.**
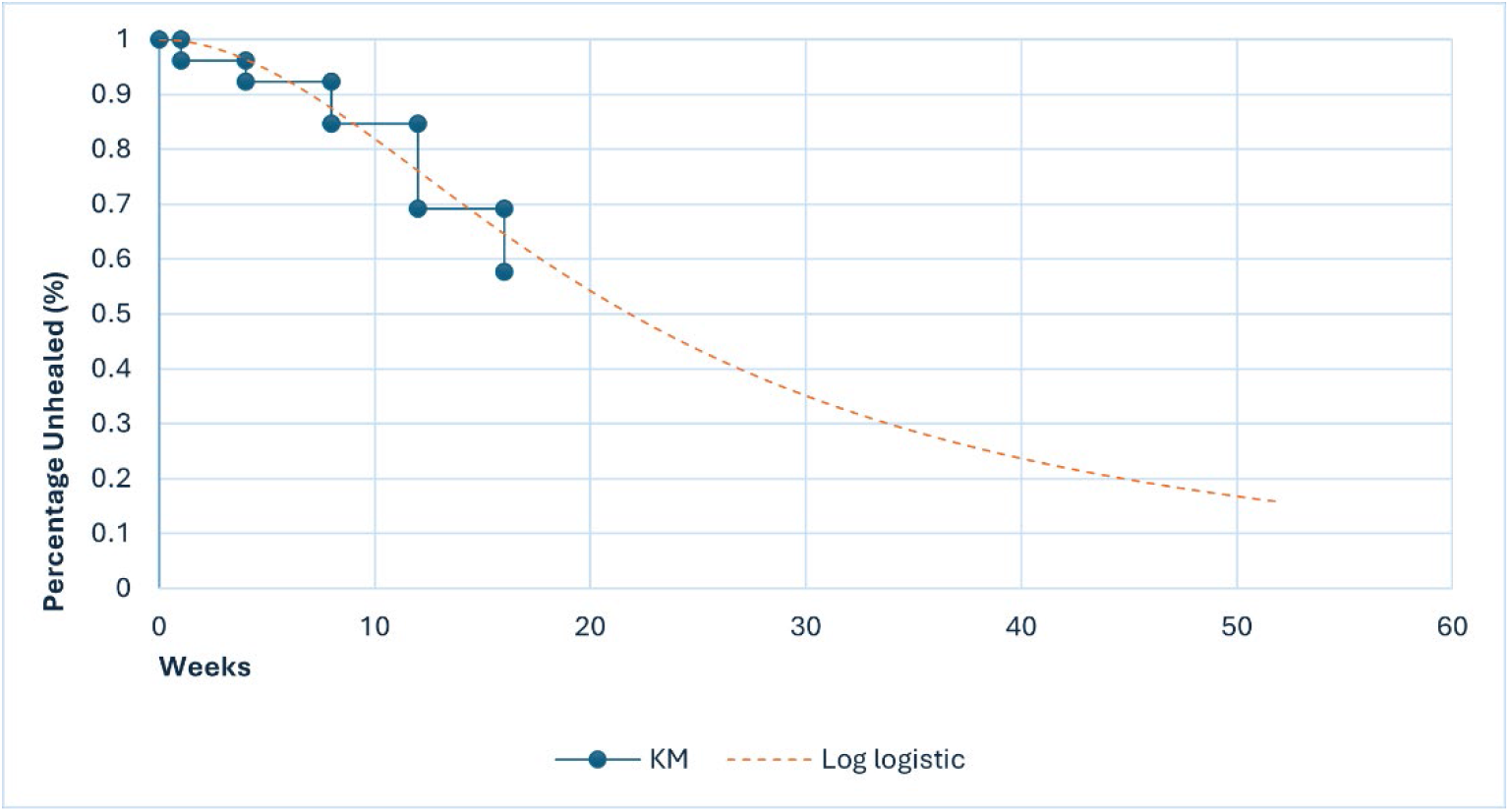
Kaplan–Meier (KM) time-to-healing analysis with parametric extrapolations for the active arm showing log logistic distribution. Active Arm, geko device 12 hrs/day plus SoC;

### 2.3 Healthcare Resource Use

We conducted a detailed examination of the resources used for delivering care, drawing insights from various studies to guide the usage of consumables. This involved an analysis focused on the control arm provided to the cohort, particularly emphasising visits to general practitioners and interactions with district nurses, including travel and administration time. Additionally, we evaluated the types of dressings and compression bandaging techniques applied in managing VLUs, as indicated in studies (4,6,14). To facilitate modelling, this data was compiled and analysed on a weekly basis. (Table 4)

For each health state within the model, we incorporated relevant healthcare resource usage estimates based on this cohort’s data. This approach enabled the calculation of the average weekly healthcare resources utilised in managing VLUs with SoC over a 12-month period. It was assumed that the NMES technology would be used exclusively in primary care, or community leg clinics settings.

District nurse visit frequency was modelled based on findings from ^**(6)**^, suggesting 2.33 to 3.5 visits per week. We adopted the lower estimate of 2.33 visits per week as a conservative approach. In light of the reduction in district nurse positions from over 9,000 to just over 4,000 within eight years leading to 2020 ^**(9)**^, any potential for resource optimisation is particularly valuable for the NHS. The NWCS indicates an average nursing visit duration of 36.2 minutes, with an additional 10 minutes for travel ^**(9)**^.

In the model, the application of the NMES technology over a two-week period required 7 units of the NMES technology (3.5 units per week), aligning with the clinical protocol of 12 hours on and 12 hours off. Therefore, each unit was calculated to provide two days of treatment. The clinical trial demonstrated that the NMES technology was used for four weeks in a 16-week period, followed by 12 weeks of SoC. It is also crucial to note that the SoC component was consistent across both treatment arms, with the NMES technology being the only variable element.

### 2.4 Utilities

Utility scores are instrumental in estimating a patient’s HRQoL in terms of quality-adjusted life-years (QALYs) gained from various interventions or services. Since HRQoL data were not recorded in the NMES technology RCT or routine clinical practice, we referred to published utility scores for VLUs (0.52 for an open VLU, 0.52 for an infected VLU, and 0.67 for a healed VLU) ^**(11)**^. It should be noted that the utility score typically attributed to VLUs does not adequately differentiate between infected and non-infected states. Despite the expectation that infected VLUs would yield a lower utility score due to increased severity, the commonly referenced studies do not reflect this distinction. In our view, this underrepresentation of the differential impact of infection on utility scores warrants attention. These scores, widely used in health technology assessments, including those by the UK National Institute for Health and Care Excellence (NICE), were sourced from UK population data specific to VLUs. Each health state in the model was assigned these utility scores, allowing for the estimation of QALYs at 12 months from treatment initiation.

### 2.5 Unit Costs

The model applied NHS unit resource costs at 2021/22 pricing (Table 1) (15,16)to the resources associated with each health state to estimate the total healthcare costs of managing a VLU with the active arm or the control arm over a 12-month period. Detailed cost calculations are provided in the appendix.

**Table 1.** NHS Unit costs for VLU delivery of both resource and consumables 2021/22.

| Resource | Unit cost (£) |
| --- | --- |
| Practice nurse consultation | 23.08 |
| General practitioner consultation | 29.00 |
| District nurse visit | 43.89 |
| Secondary dressing | 3.14 |
| Laboratory tests | 7.55 |
| Saline | 0.24 |
| Dressing pack | 0.58 |
| Wound contact layer | 0.80 |
| Foam layer | 2.58 |
| Compression bandaging 2 and 4 layers | 8.90 |
| NMES technology (per unit/pack 7) | 32.97/230.79 |

### 2.6 Model Outputs

The primary outcome measure was to demonstrate a cost saving for those patients with a chronic VLUs within the NHS. The secondary effectiveness indicators were the HRQoL of patients, quantified in terms of QALYs at 12 months from their entry into the model. Also, the probability of healing within 12 months following their introduction to the model. Employing 2021/22 cost data ^**(15)**^, we calculated the NHS expenses associated with patient management across a 12-month period from the point of entry into the model ^**(9)**^.

### 2.7 Cost-Effectiveness Analysis

To assess the cost-effectiveness of integrating the active arm compared to the control arm, we calculated the incremental cost per QALY gained. This calculation involved dividing the difference in anticipated costs between the two treatment strategies by the difference in QALYs accrued. An intervention was considered dominant if it led to a greater number of QALYs at a lower cost ^**(17)**^.

Additionally, we conducted Incremental Net Monetary Benefit (INMB) analysis ^**(18)**^. This approach quantified the economic value of the interventions by comparing their incremental costs and benefits. By calculating the net monetary benefit, we provided a comprehensive economic perspective on the effectiveness and efficiency of incorporating the NMES technology in the management of VLUs, alongside the SoC.

### 2.8 Sensitivity Analysis

Sensitivity analysis is a key tool in health economics, used to assess how changes in key assumptions or input values can affect the outcomes of a cost-effectiveness model. By varying these inputs within a defined range, it helps us understand the robustness of our conclusions and identify which factors have the most significant impact on the results. So, a probabilistic sensitivity analysis was conducted to assess the model’s inherent uncertainty. This involved performing 1,000 iterations of the model with varying input values. For each input, the standard error was assumed to be 10% around the mean, with gamma distributions applied to resource usage and costs, and beta distributions to probabilities and utilities. This process enabled the estimation of cost and QALY distributions and the likelihood of the active arm being more cost-effective than the control arm at various cost-per-QALY thresholds ^**(18)**^.

Additionally, a deterministic sensitivity analysis was carried out to examine the impact of individual parameter modifications. This involved adjusting each input and output parameter by ±20% from its base-case value. Utility scores were also varied by up to ±10% to understand potential outcome variations comprehensively ^**(17)**^. This then allowed for a greater understanding of the overall cost saving.

## 3: Results

### 3.1 Clinical Outcomes and Healthcare Costs

Before presenting the findings, to clarify the treatment groups: ‘the active arm’ refers to participants who used the geko device for 12 hours daily alongside SoC. In contrast, ‘the control arm’ consisted of participants receiving the SoC alone, as per the current NHS protocol for venous leg ulcer treatment.

The model outcomes revealed that patients treated with the active arm had a 0.84 probability of healing within 12 months, in contrast to a 0.50 probability for those receiving the control arm (Table 2). This suggests that the addition of the NMES technology to SoC alone would enhance the healing likelihood by approximately 65% over a year. Moreover, the QALY gain for patients under the active arm was higher by 0.04 QALY compared to the control arm (Table 2).

**Table 2.** Health outcomes and costs of the active arm vs control arm.

| Outcome | Active arm | Control arm |
| --- | --- | --- |
| Healed at 16 weeks | 0.42 | 0.27 |
| Healed at 12 months, probability | 0.84 | 0.50 |
| Mean time to healing, weeks | 25.3 | 37.6 |
| Mean number of QALYs per patient at 12 months | 0.60 | 0.56 |
| Mean cost per patient at 12 months, £ | 4416 | 5190 |

QALYs, quality-adjusted life-years; SoC, standard of care; Active Arm, geko device 12 hrs/day
plus SoC; Control Arm, NHS SoC only.

Additionally, the average healing time significantly reduced to 25.3 weeks with the active arm, compared to 37.6 weeks for control arm.

The estimated cost of managing a chronic VLU with the active arm was £4,416 per patient, whereas it was £5,190 per patient for the control arm (Table 3). Notably, district nurse visits constituted the bulk of the costs in SoC, accounting for up to 82%, with the remainder allocated to wound care products, such as compression bandaging and wound dressings. When using the active arm for treating an average chronic VLUs, an estimated 33% cost saving was observed in our modelling for the provision of compression bandaging and wound dressings alone. This reduction in expense brought the consumable cost down from £808.70 per patient under the control arm to £542.89 when the active arm was employed. This was mainly due to increased rate of healing. This suggests changing in the rate of healing can be measured when additional treatment interventions are used in addition to SoC alone.

**Table 3.** Cost-effectiveness analysis.

| Intervention | Total costs (£) | Total QALYs | Incremental cost saving (£) | Incremental QALYs | ICER (17) |
| --- | --- | --- | --- | --- | --- |
| Control arm | 5190 | 0.561 |  |  |  |
| Active arm | 4416 | 0.597 | -774.14 | 0.036 | Dominant |

Costs are presented in UK £.
ICER, incremental cost-effectiveness ratio; QALYs, quality-adjusted life-years; SoC, standard
of care; Active Arm, geko device 12 hrs/day plus SoC; Control Arm, NHS SoC only.

**Table 4.**
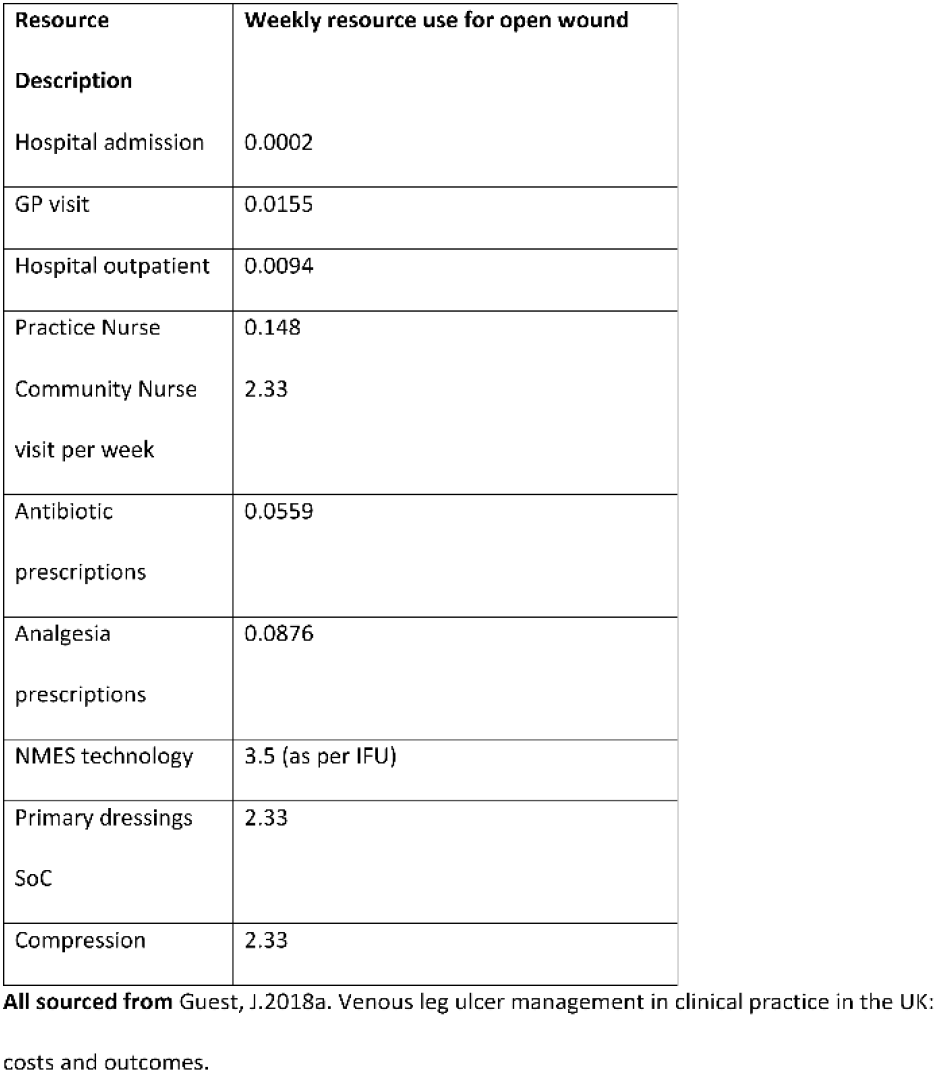
Weekly resources.

### 3.2 Cost-Effectiveness Analysis

The analysis indicated that by adding the NMES technology to SOC alone would reduce costs by £774.14 per patient over a 12-month period, with a concurrent increase of 0.04 QALYs (Table 2). Therefore, integrating the NMES technology regimen would offer a dominant treatment option for the NHS, yielding better outcomes at a reduced cost.

### 3.3 Sensitivity findings

(Figure 5) displays the incremental cost and QALY distribution between the two treatment strategies, predominantly situated in the dominant (lower cost and higher QALY) quadrant. Probabilistic sensitivity analysis suggested that, at a £20,000 per QALY threshold ^**(17)**^, there is an 85% probability that the active arm would be more cost-effective compared to control arm.

**Figure 5.**
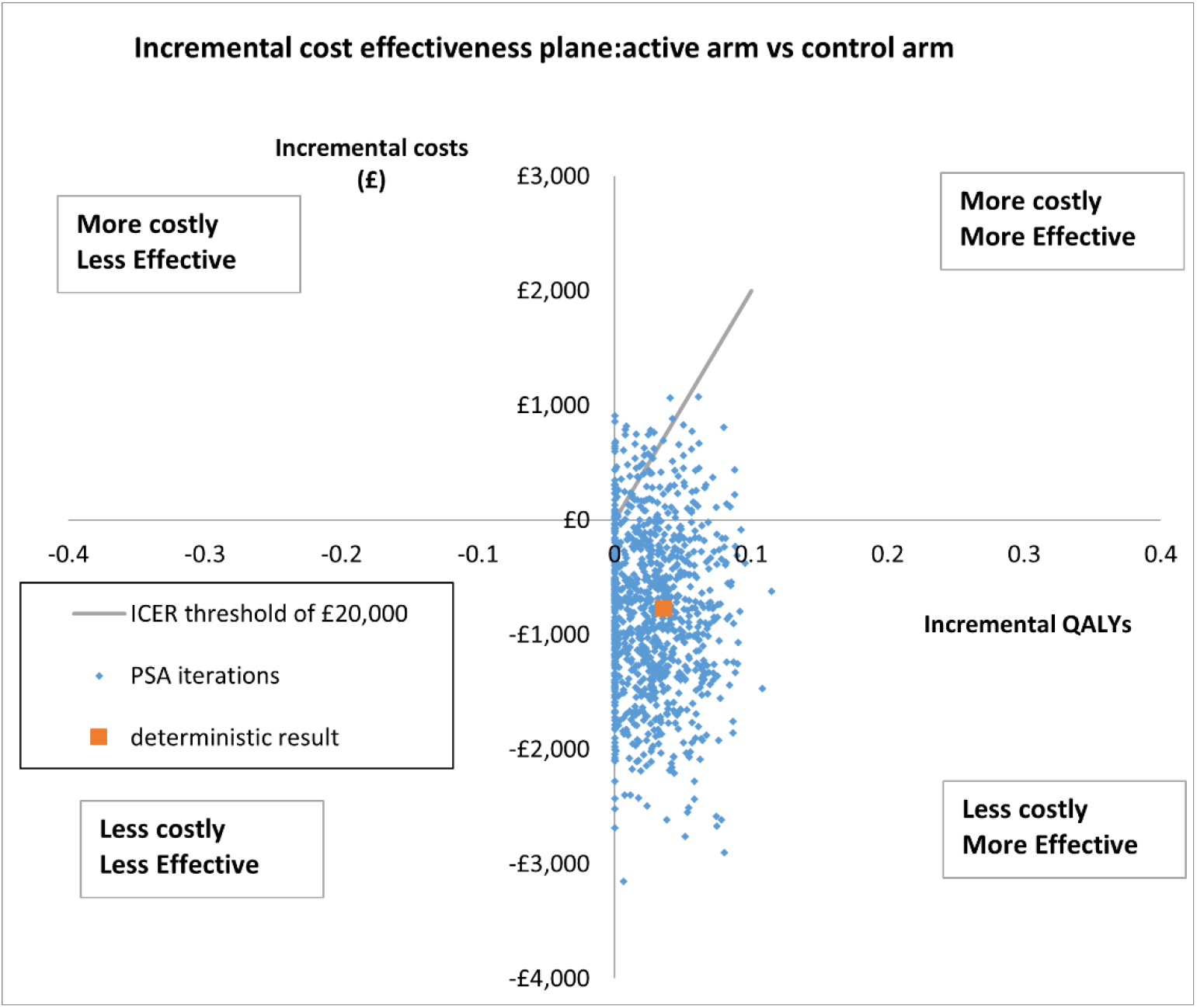
Scatterplot of the incremental cost effectiveness of the active arm vs. control arm following 1,000 iterations of the model. ICER, incremental cost-effectiveness ratio; PSA, probabilistic sensitivity analysis; QALY, quality-adjusted life-year; SoC, standard of care; Active Arm, geko device 12 hrs/day plus SoC; Control Arm, NHS SoC only.

Deterministic sensitivity analyses indicated the cost-effectiveness of the active arm was somewhat sensitive to variations in healing probability, district nurse visit frequency and utility scores. Despite these variations, the active arm consistently emerged as the dominant treatment option, even when parameter values and utility scores fluctuated by ±20% and ±10%, respectively. Therefore, the use of the NMES technology in managing chronic VLUs continues to represent a cost-effective choice, maintaining its cost-effectiveness well below the £20,000 per QALY threshold ^**(17)**^.

In the economic assessment depicted by the Cost-Effectiveness Acceptability Curve (Figure 6), the active arm shows a consistently high probability of being cost-effective across the entire range of considered QALY thresholds ^**(19)**^. This suggests a favourable balance between cost and health outcomes. Conversely, the control arm maintains a lower probability of cost-effectiveness, which gradually increases with the willingness-to-pay threshold yet remains modest. These findings indicate a potential economic advantage of the active arm, suggesting it would be a more efficient use of resources compared to the control arm. This analysis would inform decision-making processes regarding the implementation of healthcare interventions and aid clinicians and purchasers in the benefits.

**Figure 6.**
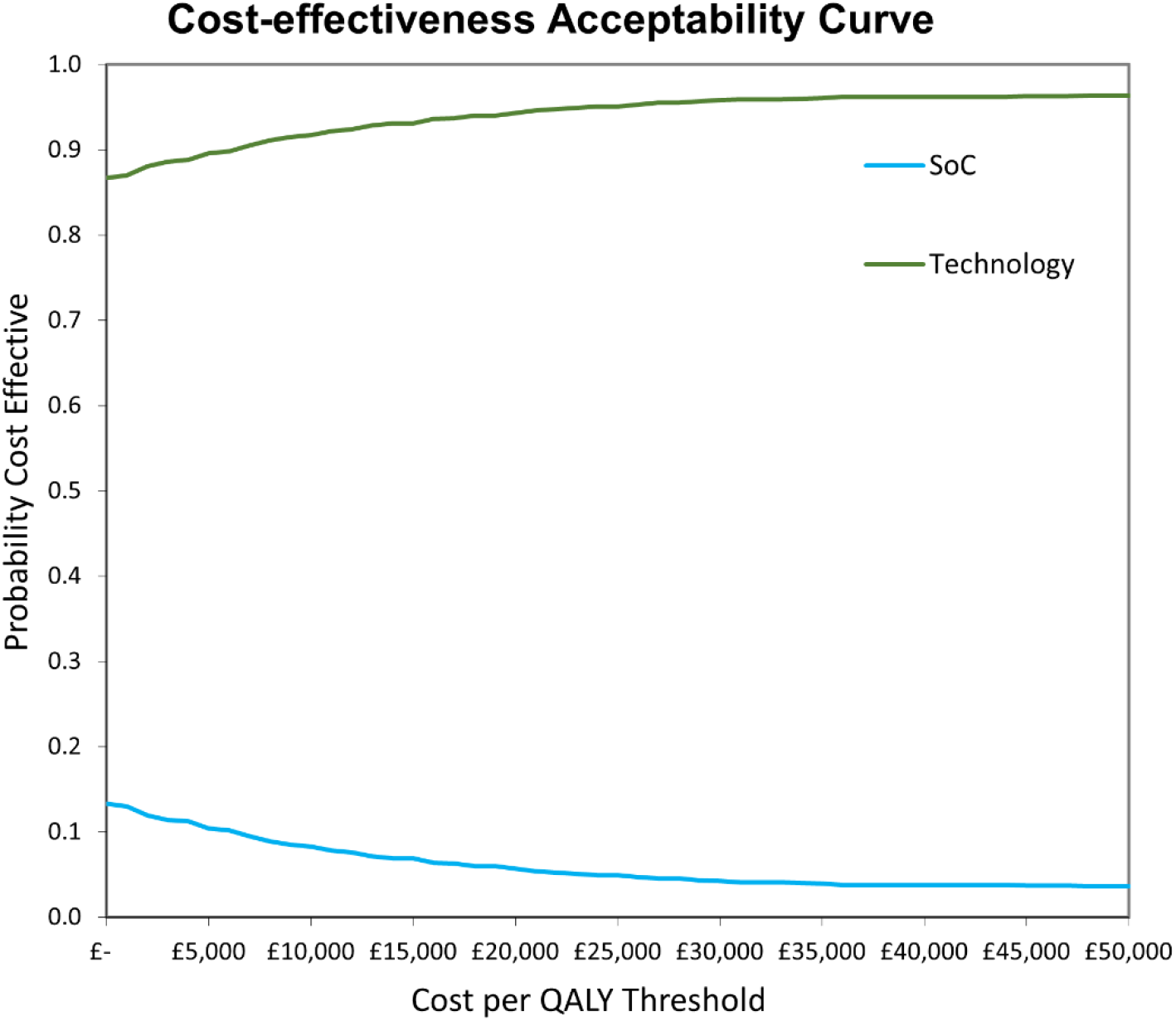
Cost-Effectiveness Acceptability Curve (CEAC) - visual representation of the Cost-Effectiveness Acceptability Curve comparing the active arm across various QALY thresholds.

### 3.4 Impact of the use of the NMES technology in your care setting

The budget impact analysis, focusing on the treatment of 100 chronic VLUs with the active arm compared to the control arm over a 12-month period, demonstrated several significant advantages:

- A 65% improvement in healing rates, translating to 34 additional patients healed per 100 treated.
- A 33% reduction in the usage of compression bandaging and wound dressings.
- A 31% decrease in the total number of district nurse visits, equating to a saving of 30 visits per patients treated.

These benefits not only offer potential system savings but also align with the NHS’s net zero targets and contribute to reducing the carbon footprint per patient.

## 4. Discussion

This study focused on the cost-effectiveness of the NMES technology in the treatment of chronic VLUs within the NHS. The findings are particularly relevant given the considerable economic and healthcare burden associated with VLUs.

As we evaluate the impact of our interventions, it is pertinent to recall the configuration of the treatment arms used in this study. ‘The active arm’ incorporated the geko device, used for 12 hours a day as an adjunct to standard care. This is in contrast to ‘the control arm’, which consisted solely of the standard care protocols currently endorsed by the NHS for managing venous leg ulcers.

To evaluate the cost-effectiveness of the active arm in treating chronic VLUs, this study employed a partitioned survival model ^**(17)**^. This model was deemed the most representative method to simulate patients’ transitions between various health states over a 12-month horizon, measured weekly. During the NMES technology RCT ^**(2)**^, the follow-up duration was 16 weeks or until healing, whichever occurred first. The control arm underwent the same 16-week follow-up. Given this, we opted to extend the model to a 12-month timeframe rather than confining it to 16 weeks. This decision was made to more accurately reflect the typical patient journey in real-world scenarios. It is important to note that beyond this 12-month period, no consideration was given to potential recurrence or additional treatments.

### 4.1 Economic Implications and Healthcare Costs

Our analysis revealed that the integration of the active arm would lead to significant savings for the NHS, both in direct treatment costs and resource utilisation. The model suggested that treating 100 chronic VLUs with the active arm, rather than the control arm, over a 12-month horizon would result in a 65% improvement in healing rates, a 33% reduction in the use of compression bandaging and wound dressings, and a 31% reduction in the total number of district nurse visits. These outcomes not only highlight the potential for more efficient resource use but also reflect a sizable monetary saving, which is crucial in the context of the NHS’s ongoing financial constraints.

### 4.2 Comparative Analysis with Existing Healthcare Costs

In comparison to previous studies and reports ^**(6)**^, such as the 2019 analysis of wound management costs in the NHS for VLUs ^**(9)**^, the potential savings with the active arm are notable and in line with other countries such as Germany with a cost of €6,905 ^**(20)**^. For instance, the annual NHS cost of managing VLUs wounds previously estimated at £3.1 billion, with a substantial portion incurred in community settings, underscores the need for more cost-effective treatments like the NMES technology ^**(9)**^. The reduction in district nurse visits and wound care product usage, as evidenced in our study, aligns well with these needs. Furthermore, the cost savings are on par with managing other significant health issues, such as obesity, which was estimated at £6.1 billion in 2014/15 and projected to rise ^**(21)**^.

### 4.3 Healthcare Management and Patient Care

The study also highlights the nurse-led nature of wound management in the NHS. The reduction in district nurse visits with the use of the active arm would not only alleviate the burden on healthcare professionals but also suggests a shift towards more efficient patient care practices. Despite this, the study reveals the need for improved management strategies, including the incorporation of new innovation such as this NMES technology, to address the high levels of resource use and costs effectively.

### 4.4 Patient Outcomes and Quality of Life

The improved healing rates and quality of life, as indicated by the QALY gains in patients treated with the control arm, demonstrate a significant clinical benefit. This aligns with the NHS’s focus on patient-centred care ^**(4)**^, emphasising not just the economic but also the human impact of effective wound management. The ability of the active arm to reduce healing time and improve patient comfort would lead to enhanced patient outcomes, potentially reducing the long-term complications associated with chronic VLUs.

### 4.5 Systemic Implications and Policy Recommendations

The findings from this study support the need for a systemic change in the approach to managing chronic wounds. The NHS, like many healthcare systems globally, faces challenges such as budget constraints and increasing demand for services. The incorporation of cost-effective and clinically beneficial innovation such as this NMES technology would be a strategic move towards more sustainable healthcare practices. It is recommended that healthcare policies and guidelines incorporate such innovative treatments, especially when they align with broader objectives like cost reduction and improved patient care.

### 4.6 Comorbidities and Care provided

Another important aspect highlighted by this study is the high prevalence of comorbidities in patients with VLUs, which aligns with previous findings ^**(14)**^. This underlines the necessity for a holistic approach to patient care that considers not only the wound itself but also the overall health and well-being of the patient. The integration of multidisciplinary care, including allied health professionals, would enhance treatment outcomes and improve the efficiency of resource use ^**(22)**^.

The management of VLUs represented a substantial financial burden on the UK NHS in 2019, with costs estimated at £3.1 billion ^**(9)**^. Of this, £2.78 billion (87%) was attributed to the management of long-standing VLUs ^**(9)**^.

In an era where healthcare demands continually escalate against the backdrop of limited resources, healthcare managers are increasingly seeking evidence of the cost-effectiveness of medical interventions to make informed decisions.

To evaluate the cost-effectiveness of the active arm in treating chronic VLUs, this study employed a partitioned survival model ^**(17)**^. This model was deemed the most representative method to simulate patients’ transitions between various health states over a 12-month period, measured weekly. During the NMES technology RCT, the follow-up duration was 16 weeks or until healing, whichever occurred first. The control arm underwent the same 16-week follow-up. Given this, we opted to extend the model to a 12-month timeframe rather than confining it to 16 weeks. This decision was made to more accurately reflect the typical patient journey in real-world scenarios. It is important to note that beyond this 12-month period, no consideration was given to potential recurrence or additional treatments.

### 4.7 Limitations

A limitation of this analysis is that it included all NHS costs and outcomes associated with wound management over the model period but did not consider the potential impact of wounds that either remained unhealed or recurred beyond the model period of 12 months. It is important to note that the analysis only considered NHS costs and resources, which are considered for the average patient. The protocol design of the RCT required any infections to be removed from the study. We were unable to review additional factors such as additional care home costs, and patient-incurred costs, which will be a key factor if patients are unable to work or perform normal activities ^**(10)**^. A cost-effectiveness assessment of the active arm in specific subgroups was not possible because the quantity of data was insufficient. A longer follow-up period would have been beneficial; however, many studies of VLUs are challenged with this issue. No patients with infection were included in the trial, as they were excluded either before or during the trial; however, we know that over 15% of patients do acquire infection while having a VLU. As a result, additional high costs have not been considered, which may have led to an underestimation of the costs and would have shown a greater cost saving for the NMES technology wound therapy.

## 5. Conclusion

This study offers an in-depth examination of the use of the NMES technology in managing VLUs within the context of the UK NHS, leading to several significant findings. The integration of the NMES technology demonstrated a marked improvement in healing rates and HRQoL for patients. Notably, the probability of healing was significantly higher in the active arm compared to control arm. This clinical benefit is coupled with considerable economic advantages, including cost savings and more efficient resource utilisation, which are critical factors in the current NHS financial climate. Findings from this study have significant implications for healthcare policy and clinical practice. They suggest that incorporating innovation such as this NMES technology in the management of chronic wounds would be a key strategy in improving patient outcomes while simultaneously addressing budgetary constraints. The potential reduction in district nurse visits and wound care product usage aligns with the NHS’s objectives of resource optimisation and enhanced patient care. Furthermore, there is a notable increase in the number of VLU patients requiring hospital admission due to unhealed or infected wounds. Given that the NHS waiting list stood at 7.5m million as of February 2024, reducing unnecessary admissions is crucial ^**(23)**^.

Chronic wounds, such as VLUs, impose a substantial burden on the healthcare system, both in terms of direct costs and the demand on healthcare services. The use of the NMES technology, as indicated by the study, offers a promising approach to mitigating these challenges, providing a cost-effective alternative to traditional treatment methods.

The current study’s findings are encouraging and pave the way for further exploration into the long-term benefits and broader applications of this NMES technology. Future research could be aimed at evaluating the sustained efficacy and wound recurrence rates, whilst investigating its effectiveness across various healthcare environments and within diverse patient demographics will also be instrumental in maximising its potential benefits.

In conclusion, the study presents strong evidence supporting the use of the NMES technology as a beneficial addition to the current treatment protocols for chronic VLUs. Its potential to enhance patient outcomes and yield significant economic benefits makes it a compelling option for inclusion in NHS wound care strategies and NHS England Long-Term Plan ^**(5)**^. The findings underscore the importance of continued innovation and adoption of cost-effective technologies in the field of healthcare, particularly in settings facing resource constraints and rising demands.

## Data Availability

All data produced in the present work are contained in the manuscript

## Acknowledgements

We would like to express our gratitude to the clinicians, healthcare staff, and patients involved in this study for their invaluable contributions. We appreciate the dedication and effort of all parties involved that made this research possible.

